# Sex Modifies the Effect of Genetic Risk Scores for Polycystic Ovary Syndrome on Metabolic Phenotypes

**DOI:** 10.1101/2021.10.23.21265391

**Authors:** Ky’Era V. Actkins, Genevieve Jean-Pierre, Melinda C. Aldrich, Digna R. Velez Edwards, Lea K. Davis

**Affiliations:** Department of Microbiology, Immunology, and Physiology, Meharry Medical College, Nashville, TN; Vanderbilt Genetics Institute, Vanderbilt University Medical Center, Nashville, TN; Division of Genetic Medicine, Department of Medicine, Vanderbilt University Medical Center, Nashville, TN; Department of Thoracic Surgery, Vanderbilt University Medical Center, Nashville, TN; Department of Biomedical Informatics, Vanderbilt University Medical Center, Nashville, TN; Vanderbilt Epidemiology Center, Institute of Medicine and Public Health, Vanderbilt University Medical Center, Nashville, TN; Division of Quantitative Sciences, Department of Obstetrics and Gynecology, Vanderbilt University Medical Center, Nashville, TN

**Keywords:** polycystic ovary syndrome, sex differences, polygenic risk scores, phenome-wide association study, electronic health records, mediation analysis

## Abstract

Females with polycystic ovary syndrome (PCOS), the most common endocrine disorder in women, have an increased risk of developing metabolic disorders such as insulin resistance, obesity, and type 2 diabetes (T2D). Furthermore, while only diagnosable in females, males with a family history of PCOS can also exhibit a poor cardiometabolic profile. Therefore, we aimed to elucidate the role of sex in the relationship between PCOS and its comorbidities by conducting bidirectional genetic risk score analyses in both sexes. We conducted a phenome-wide association study (PheWAS) using PCOS polygenic risk scores (PCOS_PRS_) to understand the pleiotropic effects of PCOS genetic liability across 1,380 medical conditions in females and males recorded in the Vanderbilt University Medical Center electronic health record (EHR) database. After adjusting for age and genetic ancestry, we found that European descent males with higher PCOS_PRS_ were significantly more likely to develop cardiovascular diseases than females at the same level of genetic risk, while females had a higher odds of developing T2D. Based on observed significant associations, we tested the relationship between PRS for comorbid conditions (e.g., T2D, body mass index, hypertension, etc.) and found that only PRS generated for BMI and T2D were associated with a PCOS diagnosis. We then further decomposed the T2D_PRS_ association with PCOS by adjusting the model for measured BMI and BMI_residual_ (enriched for the environmental contribution to BMI). Results demonstrated that genetically regulated BMI primarily accounted for the relationship between T2D_PRS_ and PCOS. This was further supported in a mediation analysis, which only revealed clinical BMI measurements, but not BMI_residual_, as a strong mediator for both sexes. Overall, our findings show that the genetic architecture of PCOS has distinct metabolic sex differences, but these associations are only apparent when PCOS_PRS_ is explicitly modeled. It is possible that these pathways are less explained by the direct genetic risk of metabolic traits than they are by the risk factors shared between them, which can be influenced by biological variables such as sex.

## Introduction

Polycystic ovary syndrome (PCOS) is a highly heritable endocrine disorder that affects 5%-21% of females of reproductive age who are typically diagnosed by having two or more of the following features under the Rotterdam criteria: polycystic ovaries, oligo- and anovulation, or hyperandrogenism [1–3]. Although Rotterdam is the most common PCOS criteria, other criteria can be used for diagnosis, including the National Institutes of Health criteria, the Androgen Excess and PCOS Criteria, or the 2018 International Evidence Based PCOS guidelines [2]. Each criterion slightly differs in requirements in an effort to cover the range of PCOS symptoms that are exhibited in patients. However, as a result of the differing diagnostic criteria, the heterogenous presentation of symptoms, and the prevalent comorbidities that reside outside of diagnostic requirements patients often spend years seeking a diagnosis or worse, may be one of the 75% of females estimated to be undiagnosed [4,5].

Many clinicians select criteria based on their perception of the most defining PCOS feature [6]. In some cases, as with the Androgen Excess and PCOS Society criteria [7], this will mean hyperandrogenism, a symptom that typically manifests as acne, hirsutism, or alopecia [8]. Androgen excess is also hypothesized to underlie many of the comorbid metabolic dysfunctions experienced by patients such as insulin resistance, obesity, metabolic syndrome, type 2 diabetes, and cardiovascular diseases (CVD). However, as previous studies have shown, the genetic risk factors present in patients with metabolic manifestations of PCOS could differ from others who have primary presentations of reproductive dysfunction [9,10].

PCOS is multifactorial and twin studies estimate heritability at 70% [11–14]. With an underlying polygenic architecture, multiple variants are hypothesized to be involved in the development of PCOS [15,16]. Furthermore, the variation of clinical features can be partially explained by ancestry informative markers, indicating potential population specific effects [17,18]. Rare variants in genes, such as *DENND1A*, have also been identified in family studies for PCOS alongside many other genetic variants identified from GWAS [19]. Despite the small effect size of individual common variants, aggregation of common risk variants together as a polygenic (or genetic) risk score (PRS) reflects the overall additive genetic liability to PCOS in individuals. This marker of disease risk is associated with PCOS diagnosis in multiple ancestries and offers many advantages to parsing out the genetic etiology of PCOS that is entangled with its comorbid presentations [11–13]. Furthermore, there is increasing evidence that a spectrum of clinical PCOS manifestations for PCOS is also correlated with the PRS for PCOS [11].

Therefore, in this study, we aimed to determine if the PRS for PCOS demonstrated pleiotropic associations with other health conditions in a hospital biobank population through a phenome-wide association study (PheWAS). By using a genetic model, we were able to determine sex-differentiated effects associated with PCOS genetic risk, revealing the impact of PCOS-associated inherited genetic variation in males, despite the fact that PCOS is only diagnosed in females. We further implemented several analyses to determine the mediating role of body mass index (BMI) on cardiometabolic comorbidities, and further identified the directionality of their effects through causal mediation analyses.

## Methods

### VUMC EHR-linked Biorepository

Vanderbilt University Medical Center (VUMC) is a tertiary care hospital in Nashville, Tennessee, with several outpatient clinics throughout Tennessee and the surrounding states offering primary and secondary care. Medical records have been electronically documented at VUMC since the early 1990s, resulting in a clinical research database of over 3 million EHRs referred to as the Synthetic Derivative [20]. EHRs include demographic information, health information documented through International Classification of Disease, Ninth Revision (ICD9) and Tenth Revision (ICD10) codes, procedural codes (CPT), clinical notes, medications, and laboratory values. This information is linked with a DNA biorepository known as BioVU. Use of EHRs and genetic data for this study was approved by the Vanderbilt University Institutional Review Board (IRB #160279).

### Genetic data

BioVU contains 94,474 individuals genotyped on the MEGA^EX^ platform [21]. The QC pipeline removed SNPs with low genotyping call rate (< 0.98) and individual subjects who were related (pi-hat > 0.2), had low call rates (< 0.98), sex discrepancies, and excessive heterozygosity (Fhet > 0.2). A principal component (PC) analysis (PCA) was performed on remaining individuals to determine genetic ancestry using FlashPCA2 [22]. BioVU genotyped samples were stratified by ancestral origin based on PCs herein referred to as the European (EUR) descent or African (AFR) descent dataset. Extended details for the quality control of these datasets have been described previously [23].

### Publicly available summary statistics

Genome-wide association study (GWAS) summary statistics were acquired for PCOS, BMI, diastolic blood pressure, systolic blood pressure, pulse, type 2 diabetes, heart failure, and coronary artery disease (i.e., all of the significant phenotypic associations observed in the PheWAS models) to further establish the strength of associations between PCOS and its comorbidities through subsequent analyses. Each GWAS was selected based on public availability, sample size, and sample diversity.

The Genetic Investigation of ANthropometric Traits (GIANT) consortium body mass index (BMI) summary statistics included 339,224 individuals of European and non-European descent from over 125 studies. These BMI summary statistics were used as a proxy for obesity summary statistics [24]. Blood pressure traits (diastolic, systolic, and pulse) and type 2 diabetes (T2D) summary statistics were obtained from the Million Veterans Program (MVP), a large biobank consortium effort that houses biobank data from various sites in the Department of Veterans Affairs health system [25]. Blood pressure traits were generated from a trans-ethnic sample of over 750,000 individuals from MVP [26]. T2D summary statistics were generated from a meta-analysis using data from 1.4 million participants in various biobanks and consortia groups [27]. Heart failure summary statistics were collected from 47,309 cases and 930,014 controls of European ancestry across nine studies in the Heart Failure Molecular Epidemiology for Therapeutic Targets (HERMES) consortium as a proxy for heart disease [28]. Finally, coronary artery disease (CAD) datasets generated from the Coronary Artery Disease Genome-wide Replication and Meta-analysis plus The Coronary Artery Disease (CARDIoGRAMplusC4D) consortium were used as the genetic measurement for the coronary atherosclerosis phenotype [29]. This meta-analysis assembled 60,801 cases and 123,504 controls of multiple ancestries across forty-eight study sites.

### Statistical analysis

#### Generation of polygenic risk scores (PRS)

PCOS_PRS_ were calculated with PRS-CS software using the weighted sums of the risk allele effects as reported in the summary statistics from the Day et al. GWAS of PCOS and applying a Bayesian continuous shrinkage parameter select SNP features and to model linkage disequilibrium [16,30]. The details of these methods have been previously described elsewhere [11]. We calculated PCOS_PRS_ for both EUR and AFR BioVU genotyped ancestry samples which were previously shown to be associated with a PCOS diagnosis defined by a coded strict PCOS definition in our previously published EHR-based algorithm [11].

#### Phenome-wide association study (PheWAS)

Next, we were interested in identifying the pleiotropic effects of the genetic susceptibility to PCOS on the medical phenome. Therefore, in a PheWAS framework we analyzed the effects of PCOS_PRS_ across 1,380 medical conditions. This analysis was first performed in a female sample for our EUR and AFR ancestry datasets to validate the PRS and identify potential pleiotropic relationships (**Supplementary Figures 1-5**). Although males cannot be diagnosed with PCOS, they still harbor genetic risk for PCOS. Thus, we extended the analysis to males to examine sex differences. Finally, we performed a sex-combined PheWAS to increase statistical power. In the sex-combined logistic regression model, covariates included median age of individuals medical record, sex, and the first ten principal components. In the sex-stratified models, covariates included median age and the first ten PCs.

#### Interaction analysis

For each phenotype with evidence of a significant main effect of PCOS_PRS_ in either sex, we tested for two-way interactions (sex * PCOS_PRS_) to determine which of the significant sex stratified PheWAS associations were influenced by biological effects of sex. Selected phenotypes of interest that met the phenome-wide false discovery rate (q < 0.05) were tested for interactions (N = 10). For these interaction analyses, sex-combined models included the main effects of PCOS_PRS_, median age of individuals medical record, sex, and the top ten PCs.

#### Sensitivity analyses

Several sensitivity analyses were performed to assess the robustness of the significant phenome-wide findings. First, BMI is strongly correlated with both PCOS and its comorbidities, and thus, can influence the strength of the results [31]. Therefore, we adjusted each model for BMI (median measurement across an individual’s EHR) to observe whether the significant phenotypes associated with PCOS_PRS_ were independent of obesity-related effects. In addition to adjusting for BMI, the model was adjusted for median age, sex (only in the sex-combined sample), and the top ten PCs.

Next, we evaluated which phenotypes were dependent on a PCOS diagnosis in females by accounting for PCOS diagnosis [11]. This analysis allowed us to identify true pleiotropic associations and provided insight into which phenotypes were exclusively correlated with genetic risk, even in the absence of PCOS.

Finally, given that BMI has strong contributions from both genetic and environmental sources of variance, we revisited the previous PheWAS analyses to specifically account for the environmental contribution of BMI. To do this, we calculated the residuals of median BMI adjusted for BMI_PRS_ (residuals(medianBMI ∼ BMI_PRS_)), herein referred to as BMI_residual_. BMI_residual_ were then used in a subsequent sensitivity analysis of the previously described PheWAS.

#### Genetic correlation

Linkage Disequilibrium Score Regression (LDSC) was used to calculate genetic correlation, an estimate of genetic similarity, between traits [32]. LDSC only utilizes GWAS summary statistics and is not sensitive to sample overlap, which may be present across the publicly available GWAS datasets used in this study. This method utilizes the effect estimate of each SNP and accounts for the effects of SNPs in linkage disequilibrium based on the GWAS reference population. The European reference panel was used for all analyses based on the demographic majority of each of the GWAS samples.

#### Logistic regression models

Public GWAS datasets were used to generate PRS for phenome-wide significant phenotypes identified in the PheWAS. PRS were then used as the independent variable in a logistic regression model against PCOS diagnosis as the dependent variable. BioVU genotyped datasets were filtered to females and contained 361 PCOS cases and 29,035 controls in the EUR dataset and 189 PCOS cases and 9,229 controls in the AFR dataset. Selection of PCOS cases and controls have been described elsewhere [11]. In brief, cases required PCOS billing codes and no exclusion codes. Controls excluded individuals who had any inclusion or exclusion codes [11]. All models were adjusted for median age of the individuals medical record, and the top ten PCs for each ancestry. Median BMI was included as a covariate in the sensitivity analysis.

Lastly, we adjusted for BMI_residual_ in the logistic regression models in place of BMI. Next, we performed a sensitivity analysis to test the effect of BMI_residual_ and BMI_PRS_ separately on PCOS diagnosis. A multiple testing correction of p < 7.35e-04 (0.05/68) was implemented to account for all statistical tests. All statistical analyses were done using R 3.6.0.

#### Mediation analysis

To test whether BMI or BMI_residual_ mediates the pleiotropic relationships between PCOS and associated cardiometabolic conditions, a mediation analysis was employed using the *mediation* R package for both AFR and EUR genotyped samples [33]. For the first set of mediation analyses, we modeled BMI_PRS_ and BMI_residual_ as exposures, T2D diagnosis as the mediator, and PCOS case status as the outcome. Another reciprocal model was tested with the same exposures, but with PCOS diagnosis as the mediator and T2D as the outcome. This analysis tested the hypothesis that the risk conferred by BMI on one phenotype (e.g., PCOS) in turn increases risk for another other phenotype (e.g., T2D). We restricted this analysis to the female population. All models were further adjusted for median age and the top ten PCs.

In our second set of mediation analyses, we tested whether genetic risk for PCOS or T2D also influenced BMI_residual_ through the manifestation of either clinical condition. For example, we modeled PCOSPRS as the exposure variable, T2D diagnosis as the mediator and BMI_residual_ as the outcome. Finally, in a female only analysis, T2DPRS was modeled the exposure variable, PCOS diagnosis was included as the mediator, and BMI_residual_ again was modeled as the outcome.

Finally, for our last set of mediation analyses, we examined the mediating effect of both measured BMI and BMI_residual_ on the relationship between PCOS_PRS_ (exposure) and the diagnosis of hypertension and hypertensive heart disease (i.e., outcomes that demonstrated evidence of significant interactions with sex). T2D was also tested because it was significantly associated with PCOS_PRS_ in both sexes. PCOS_PRS_ was modeled as the exposure variable and T2D (phecode = 270.2) hypertension (phecode = 411), and hypertensive heart disease (phecode = 401.21) were modeled as the outcome variables. Models were considered significant when the average direct effect (ADE) and average causal mediation effect (ACME) both passed p < 7.35e-04. All mediation analyses were performed with 1,000 bootstrap simulations to estimate the confidence intervals and determine an empirical p-value.

## Results

### PCOS_PRS_ PheWAS results

The PCOS_PRS_ PheWAS was applied to 66,903 EUR individuals from BioVU. When restricted to females (N = 37,240), two phenotypes were significantly associated with PCOS_PRS_ (Bonferroni corrected p-value = 4.56e-05) (**Figure 1a**): T2D (odds ratio [OR] = 1.10, 95% confidence interval [CI] = 1.06-1.14, p = 5.54e-07) and diabetes mellitus (OR = 1.09, 95% CI = 1.05-1.13, p = 2.20e-06). When the analysis was restricted to males (N = 29,663; **Figure 1b**), hypertensive heart disease was significant (OR = 1.15, 95% CI = 1.08-1.23, p = 2.07e-05) alongside a cluster of nominally (p < 0.05) associated cardiovascular phenotypes with similar effect sizes (OR = 1.06 to 1.08). This included hypertension (OR =1.06, 95% CI = 1.03-1.10, p=1.30e-04), essential hypertension (OR = 1.06, 95% CI = 1.03-1.10, p = 2.49e-04), and coronary atherosclerosis (OR = 1.06, 95% CI =1.02-1.10, p=1.40e-03). T2D (OR=1.07, 95% CI = 1.03-1.11, p=7.04e-04) and several other endocrine phenotypes were also modestly associated with PCOS_PRS_ in males.

**Figure 1.**
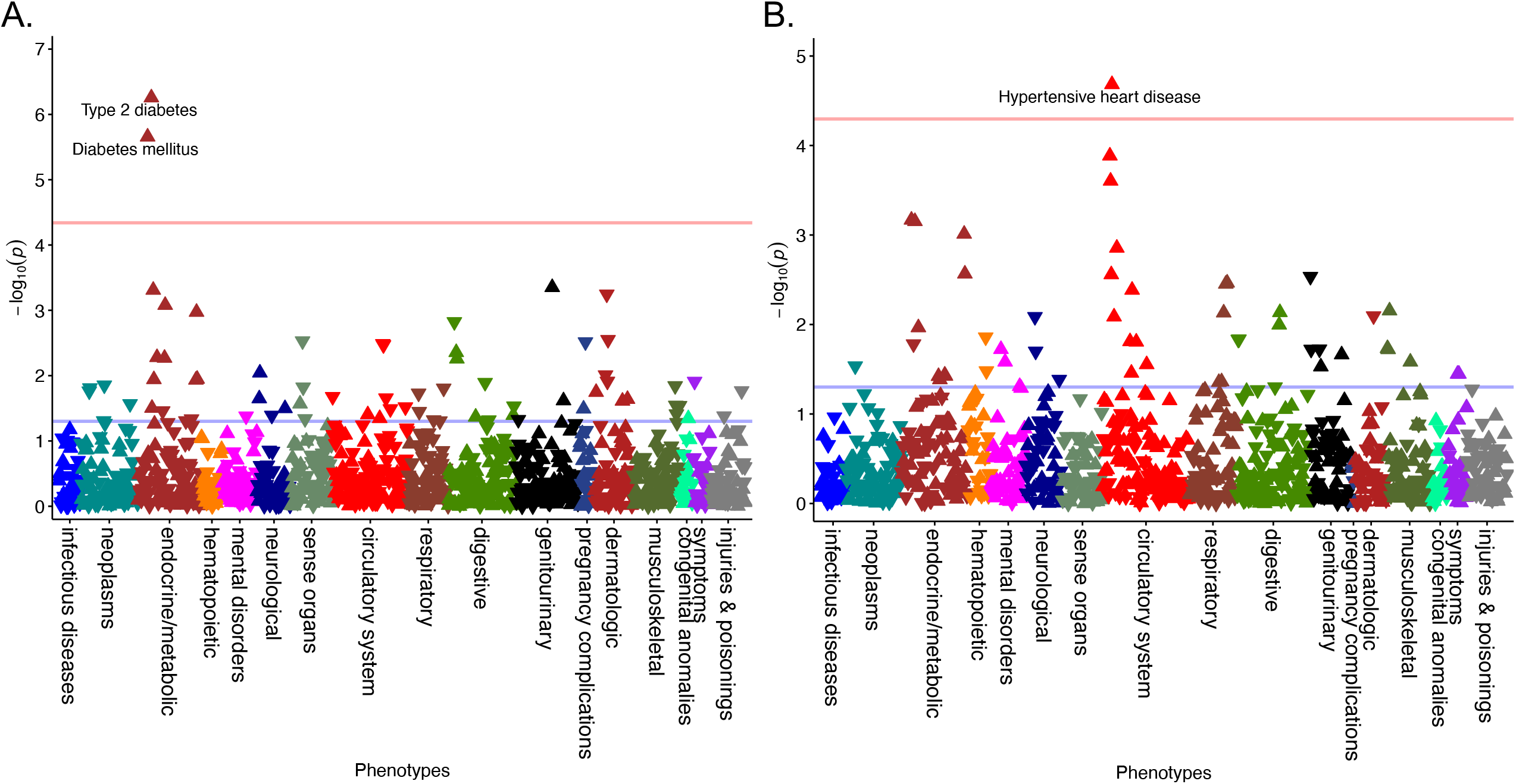
Metabolic Associations Vary by Sex for Genetically High-Risk Individuals. PCOS_PRS_ phenome-wide association study results are displayed in the Manhattan plots for (A) females and (B) males. The red line represents the Bonferroni correction of P = 4.56e-05 and P = 5.07e-05, respectively. Arrows in the upward direction represent increased risk.

In the EUR sex-combined model, four phenotypes were significantly associated with PCOS_PRS_ (**Figure 2**). This included T2D and diabetes mellitus as the top associations with the same OR of 1.08 (T2D 95% CI = 1.06-1.11, p = 3.15e-09; diabetes mellitus 95% CI = 1.05-1.11, p = 1.03e-08). Following was obesity (OR = 1.07, 95% CI = 1.04-1.11, p = 9.68e-06) and hypertensive heart disease (OR = 1.11, 95% CI = 1.06-1.16, p = 3.57e-05). Three cardiovascular diseases were also nominally associated with PCOS_PRS_, presumably due to the addition of males. PCOS_PRS_ yielded an OR of 1.21 (95% CI = 1.08-1.36, p = 7.82e-04) for an association with polycystic ovaries, falling just short of phenome-wide significance.

**Figure 2.**
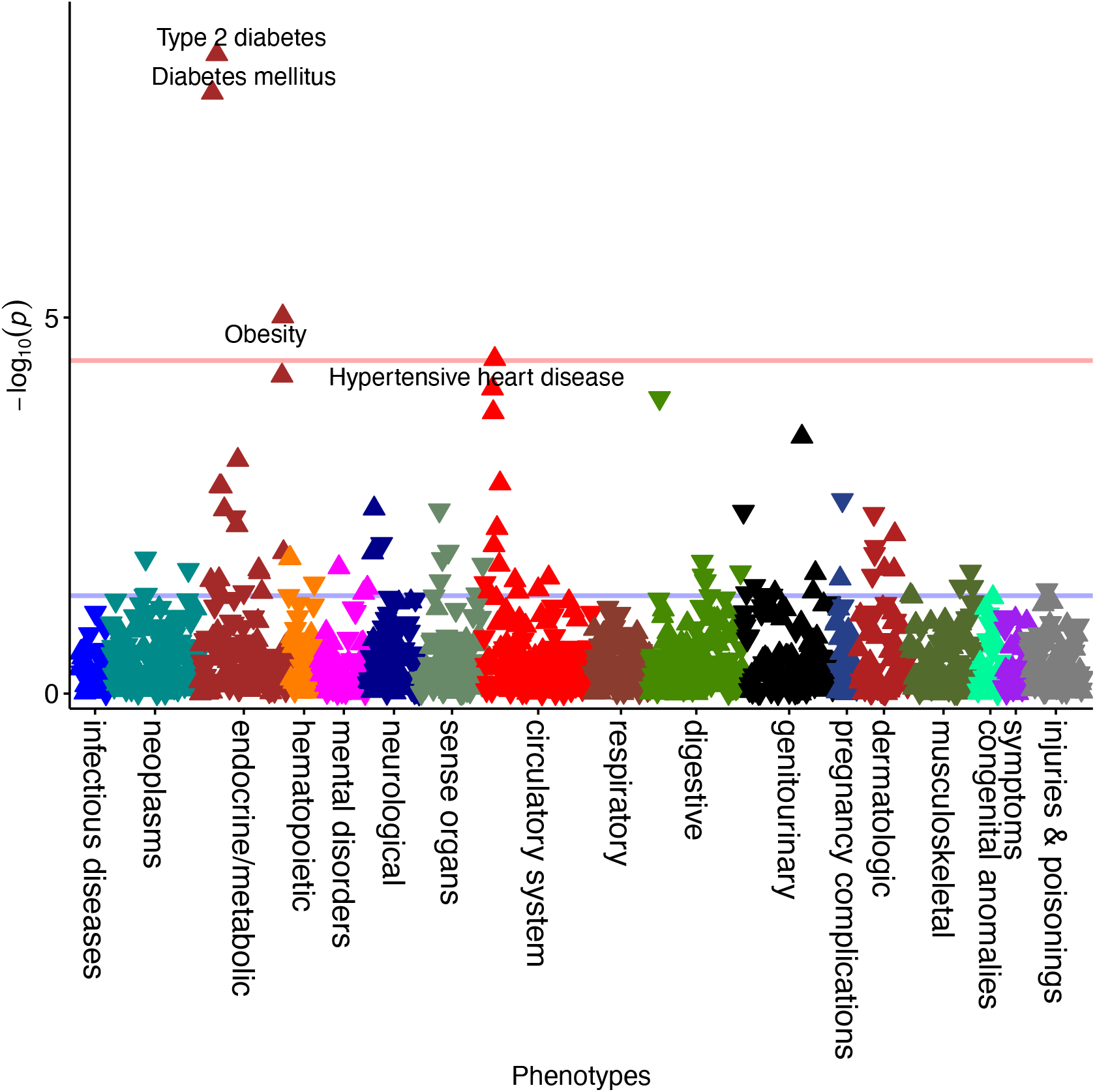
Sex Combined PCOS_PRS_ PheWAS Analysis Increases Detection of Associations. The phenome-wide association study (PheWAS) was performed on the full dataset (females and males) to increase power. Significant results that passed Bonferroni correction (P = 3.73e-05) are annotated in the Manhattan plot.

No associations passed Bonferroni correction in the AFR ancestry analysis, which included a total of 12,383 individuals (**Supplementary Figure 1**).

### Interaction analysis

The sex interaction analysis demonstrated that males with a high PCOS_PRS_ were (interaction p = 7.24e-03) more likely to be diagnosed with hypertension (OR_Males_ =1.06, 95% CI = 1.03-1.10, p =1.30e-04; OR_Females_ = 1.03, 95% CI = 1.00-1.06, p = 0.07), essential hypertension (interaction p=7.71e-03, OR_Males_ = 1.06, 95% CI = 1.03-1.10, p = 2.49e-04; OR_Females_ = 1.03, 95% CI = 1.00-1.06, p = 0.08), and hypertensive heart disease (interaction p=0.01, OR_Males_ = 1.15, 95% CI = 1.08-1.23, p = 2.07e-05; OR_Females_ = 1.05, 95% CI = 0.98-1.13, p = 0.16) than females with the same PCOS_PRS_ (**Table 1**). These sex differences were also observed when calculating the prevalence for each trait by decile of PCOS_PRS_, which only showed an upwards trend across deciles for traits with a significant sex effect (**Figure 3**). Together, these results showed that sex is an important modifier of PCOS genetic risk.

**Table 1.**
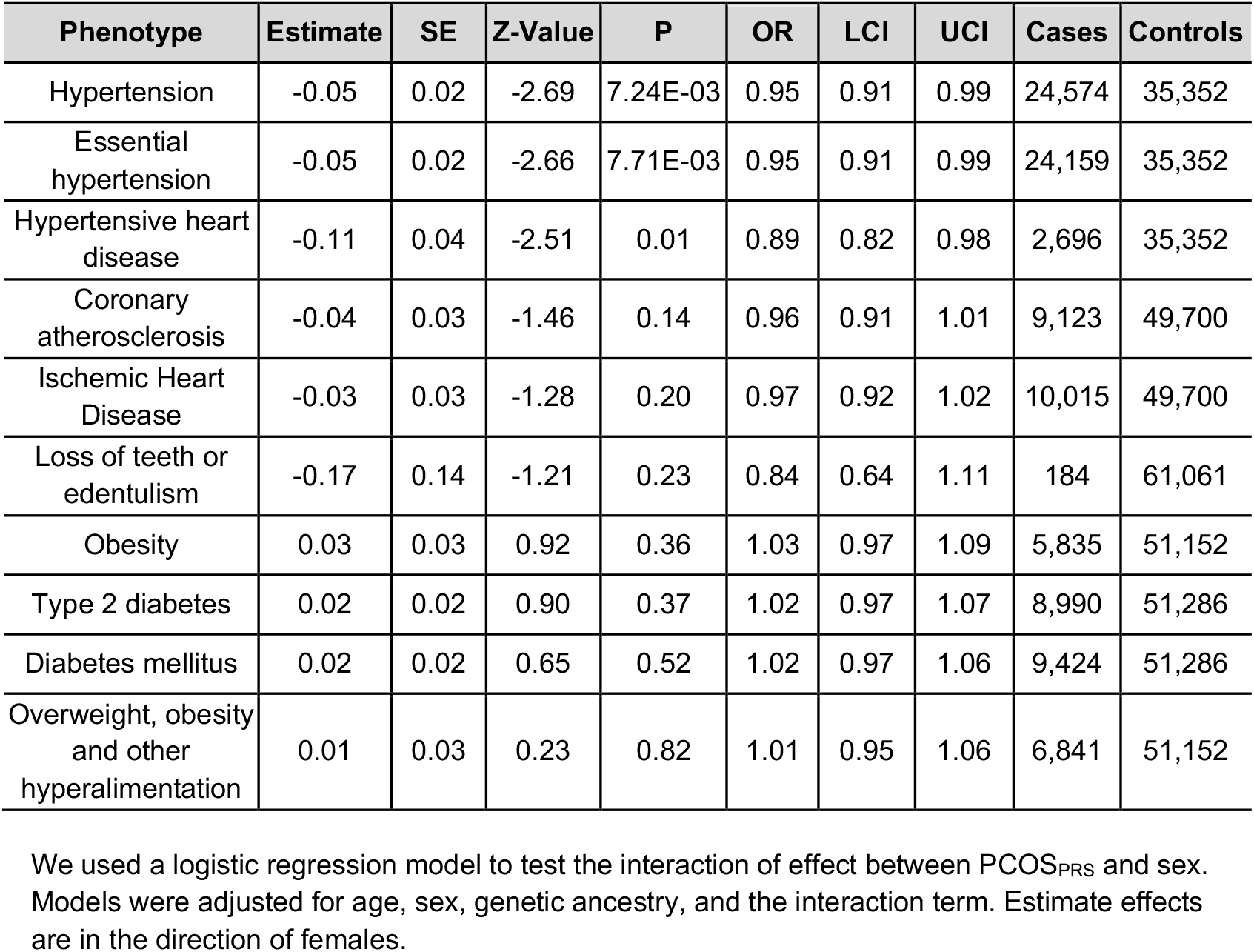
Hypertension is Significantly Modified by Sex for PCOS_PRS_.

**Figure 3.**
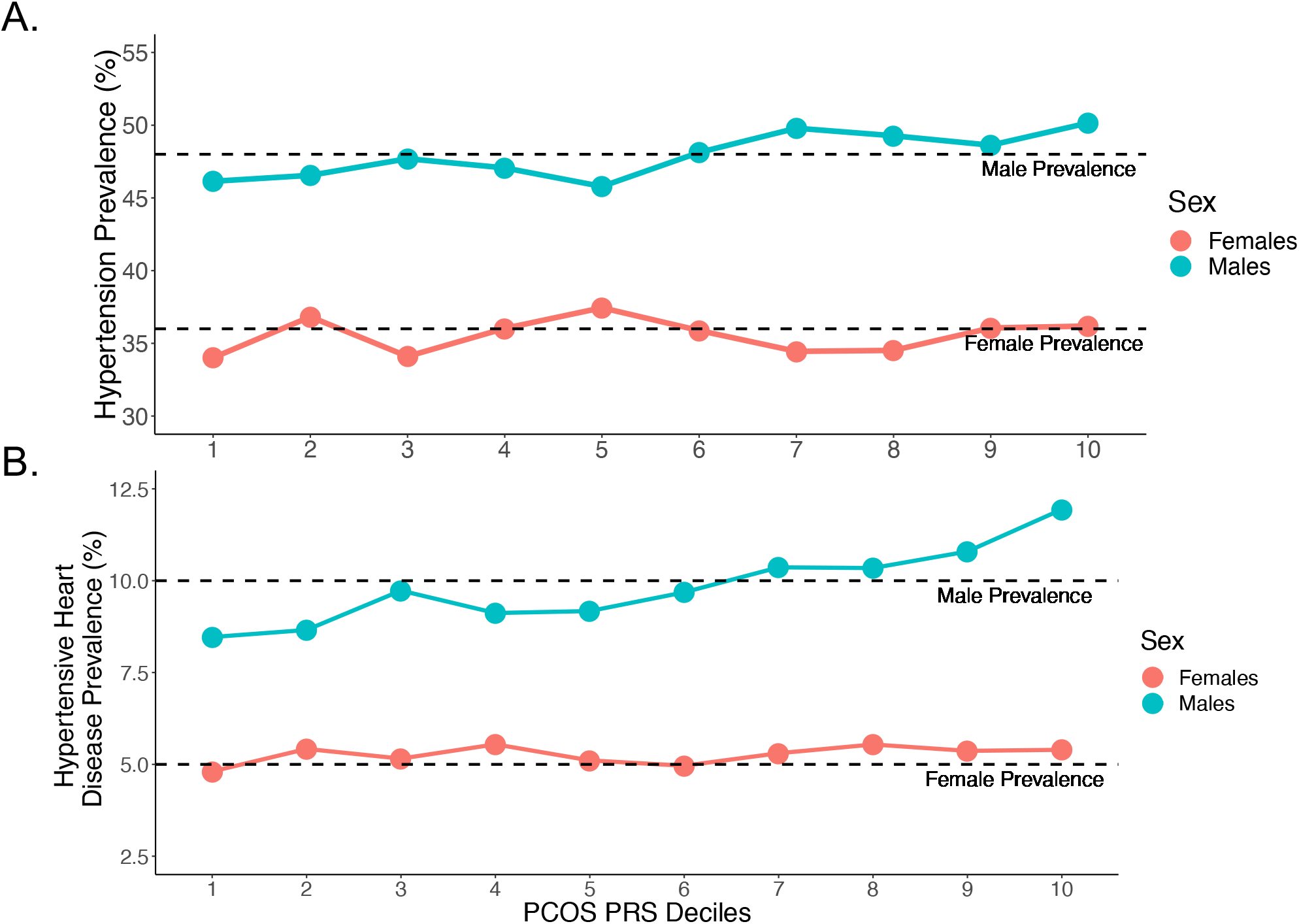
The Prevalence of Hypertensive Metabolic Traits Differs Across the PCOS_PRS_ Strata. (A) Top Figure: Hypertension, (B) Bottom Figure: Hypertensive Heart Disease. The prevalence of the significant phenotypes from the sex interaction analysis are shown. The prevalence for hypertension in our dataset was 48% for males and 36% for females. The prevalence for hypertensive heart disease was 10% for males and 5% for females.

### Sensitivity analyses adjusting for BMI and PCOS case status

In our first set of PheWAS sensitivity analyses for the EUR population, we adjusted for median BMI in all of the PRS-PheWAS models. There were no surviving associations in the sex-combined model or stratified analyses, suggesting BMI may mediate the pleiotropic effects of PCOS_PRS_ (**Supplementary Figures 2a and 3**). In a separate sensitivity analysis (female only) we found that after adjusting for PCOS diagnosis, females with a high PCOS_PRS_ still demonstrated a significant positive phenome-wide association with T2D and diabetes mellitus (**Supplementary Figure 2b**).

### Genetic correlation results

We found that BMI (r_g_ = 43.32%, p = 8.77e-19) and T2D (r_g_ = 29.26%, p = 3.25e-09) had the strongest genetic correlation with PCOS (**Table 2**). Heart failure (r_g_ = 26.51%, p = 0.0025) and pulse pressure (r_g_ = 13.46%, p = 0.011) were also modestly significantly with PCOS. CAD bordered the significance threshold, but systolic and diastolic blood pressure were not significantly genetically correlated with PCOS.

**Table 2.** PCOS Shares Genetic Architecture with Cardiometabolic Traits.

| Traits | Genetic Correlation | SE | Z | P |
| --- | --- | --- | --- | --- |
| Diastolic Blood Pressure | -3.60% | 0.07 | -0.54 | 0.59 |
| Systolic Blood Pressure | 9.33% | 0.05 | 1.79 | 0.07 |
| Pulse Pressure | 13.46% | 0.05 | 2.54 | 0.01 |
| Type 2 Diabetes | 29.26% | 0.05 | 5.92 | 3.25e-09 |
| Heart Failure | 26.51% | 0.09 | 3.02 | 0.003 |
| Coronary Artery Diseases | 17.35% | 0.09 | 1.95 | 0.05 |
| Body Mass Index | 43.32% | 0.05 | 8.85 | 8.77e-19 |

### Cardiometabolic PRS Analysis of PCOS diagnosis

Outside of T2D and BMI, none of the PRS built for CAD, heart failure, or blood pressure were significantly associated with a PCOS diagnosis in females of either EUR or AFR descent (**Figure 4**).

**Figure 4.**
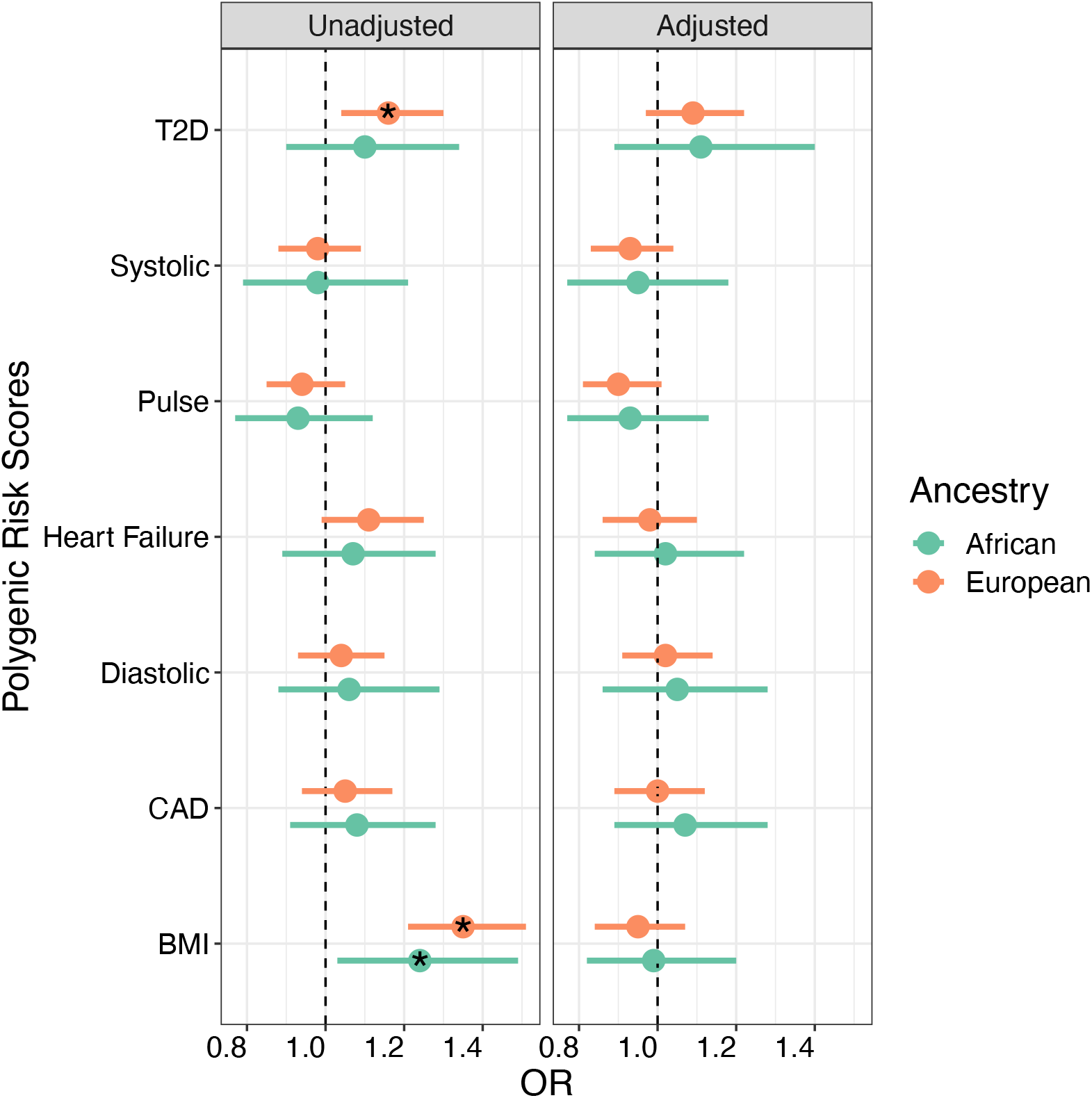
T2D_PRS_ and BMI_PRS_ are Associated with PCOS Case Status. The results from the logistic regression analysis between PCOS diagnosis and polygenic risk scores (PRS) generated for significant metabolic traits from the PheWAS analysis are displayed in the forest plot. One model was unadjusted for body mass index (unadjusted) and the other model was adjusted for body mass index (adjusted). * indicates P < 0.05

BMI_PRS_ was positively associated with PCOS diagnosis for both EUR and AFR ancestry populations (p_EUR_ = 3.71e-08, p_AFR_ = 0.02), but not independently of clinically measured BMI values. T2D_PRS_ in the EUR dataset yielded similar results with PCOS diagnosis, again losing significance upon addition of a BMI covariate in the model (OR_unadjusted_ = 1.16, 95% CI = 1.04-1.30, p = 0.007; OR_adjusted_ = 1.09, 95% CI = 0.97-1.22, p = 0.16). To determine whether this reduction in effect size was due to the genetic correlation between T2D and BMI, we also tested a model in which T2D_PRS_ was adjusted for BMI_residual_ (i.e., variance remaining in BMI after removing variance due to BMI_PRS_) instead of BMI. This model indeed recovered the original association between T2D_PRS_ (OR_EUR_ = 1.18, 95% CI = 1.05-1.32, p_EUR_ = 0.005) and PCOS diagnosis for the EUR population, suggesting that *genetically predicted* BMI mediates the association between T2D_PRS_ and PCOS diagnosis (**Figure 5**).

**Figure 5.**
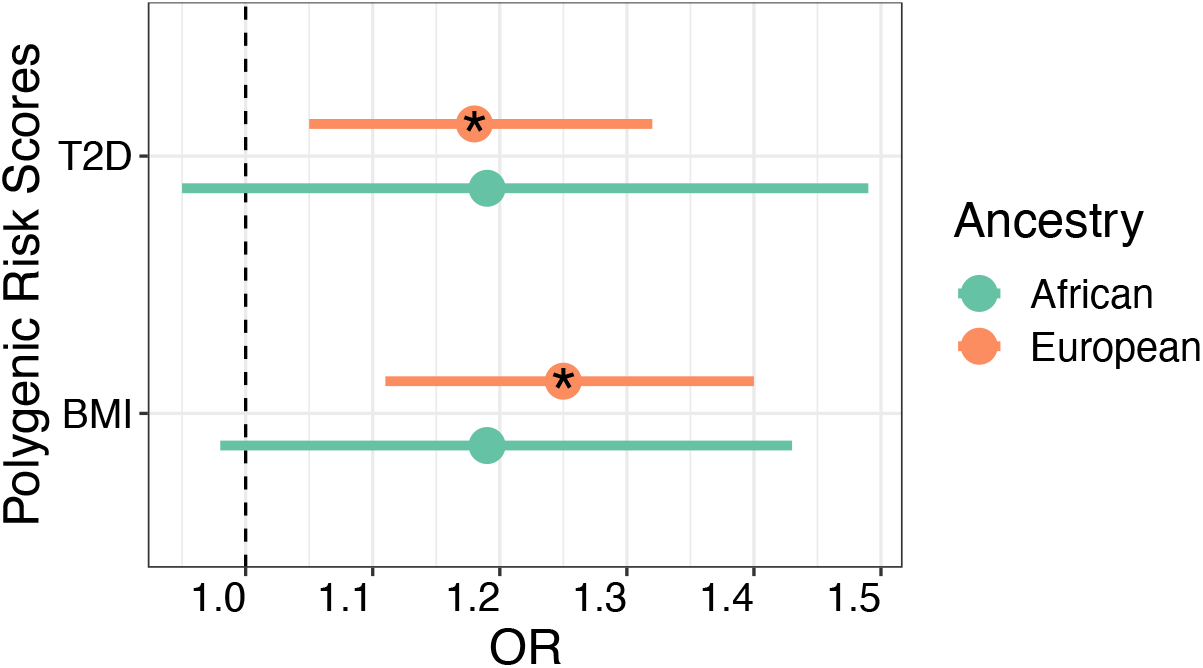
T2D_PRS_ is Associated with PCOS After Accounting for BMI_residual_. The logistic regression model between T2D_PRS_ and PCOS diagnosis was covaried for age, the top ten principal components for ancestry, and BMI_residual_. BMI_residual_ represents BMI after the removal of BMI genetic variance from the variable. * indicates P < 0.05

### Mediation analysis in females with BMI as an exposure variable

To further quantify the degree to which BMI influenced the shared genetic pathways between PCOS and T2D, a mediation analysis was performed with BMI_PRS_ and BMI_residual_ as the exposure variables, PCOS diagnosis as the outcome variable, and T2D as the mediator. The reciprocal model was then tested with T2D diagnosis as the outcome and PCOS as the mediator (**Table 3**). We found that both phenotypes acted as significant mediators when BMI_PRS_ and BMI_residual_ were exposure variables. PCOS diagnosis had a stronger mediating effect on T2D when BMI_residual_ was the exposure variable compared to BMI_PRS_ (13.6% vs 7.1%). However, the opposite was true for T2D, which mediated more of risk conferred by BMI_PRS_ than BMI_residual_ on the PCOS outcome (9% vs 2.1%).

**Table 3.** BMI Genetic Variance is a Causal Risk Factor for PCOS and T2D.

| Exposure | Mediator | Outcome | ADE<br>$\beta$ (95% CI) | ACME<br>$\beta$ (95% CI) | Proportion Mediated<br>(95% CI) |
| --- | --- | --- | --- | --- | --- |
| BMI<sub>PRS</sub> | PCOS | T2D | 0.025<br>(0.014-0.037)<br>$P = <2e-16$ | 0.002<br>(6.92E-04-0.004)<br>$P = <2e-16$ | 0.071<br>(0.025-0.144)<br>$P = <2e-16$ |
| | T2D | PCOS | 0.025<br>(0.013-0.037)<br>$P = <2e-16$ | 0.002<br>(0.001-0.004)<br>$P = <2e-16$ | 0.090<br>(0.039-0.191)<br>$P = <2e-16$ |
| BMI<sub>residual</sub> | PCOS | T2D | 0.037<br>(0.023-0.052)<br>$P = <2e-16$ | 0.006<br>(0.002-0.010)<br>$P = <2e-16$ | 0.136<br>(0.049-0.254)<br>$P = <2e-16$ |
| | T2D | PCOS | 0.127<br>(0.104-0.150)<br>$P = <2e-16$ | 0.003<br>(8.12-04- 0.005)<br>$P = 0.004$ | 0.021<br>(0.006-0.042)<br>$P = 0.004$ |

ADE = average direct effect, ACME = average causal mediation effect, CI = confidence interval

### PheWAS sensitivity analysis with BMI_*residual*_

We revisited the PheWAS analysis in light of the findings for BMI_residual_. By covarying for BMI_residual_, (e.g., environmental estimate of BMI), we observed almost no difference from the original results in our female sample (**Supplementary Figure 4a**). T2D remained significant in females (OR =1.10, 95% CI = 1.06-1.15, p = 1.69e-06), as did diabetes mellitus (OR =1.09, 95% CI = 1.05-1.13, p = 9.91e-06). These associations also appeared in the combined dataset alongside obesity (**Supplementary Figure 5**). Although none of the phenotypes passed Bonferroni correction for males, many of the associations improved in estimate effects (**Supplementary Figure 4b**).

### Mediation analysis in the sex-combined and sex-stratified samples

Upon finding BMI_residual_ as a significant risk exposure for T2D and PCOS in the first mediation analysis, but not a strong confounder in the PCOS_PRS_ PheWAS analysis, we tested whether the genetic risk conferred by one condition (e.g., T2D_PRS_) was mediated by the clinical manifestation of the other (e.g., PCOS diagnosis) when examining BMI_residual_ as the outcome. Here we observed a modest association that showed PCOS diagnosis mediated 33% of the risk conferred by T2D_PRS_ on BMI_residual_ whereas T2D diagnosis did not explain any of the variance in BMI_residual_ due to PCOS_PRS_ (**Table 4**).

**Table 4.** Genetic effects of T2D Contributes to Environmental BMI.

| Sex | Exposure | Mediator | Outcome | ADE<br>$\beta$ (95% CI) | ACME<br>$\beta$ (95% CI) | Proportion<br>Mediated<br>(95% CI) |
| --- | --- | --- | --- | --- | --- | --- |
| Sex-Combined | PCOS<sub>PRS</sub> | T2D | BMI<sub>residual</sub> | 0.041<br>( -0.055-0.147)<br>*P* = 0.44 | 0.059<br>(0.033-0.088)<br>*P* = <2e-16 | 0.589<br>(-1.112-3.248)<br>*P* = 0.06 |
| Females |  |  |  | 0.024<br>(-0.102-0.172)<br>*P* = 0.78 | 0.059<br>(0.041-0.121)<br>*P* = <2e-16 | 0.652<br>(-4.638-6.128)<br>*P* = 0.18 |
| Males |  |  |  | 0.040<br>(-0.101-0.176)<br>*P* = 0.58 | 0.038<br>(0.004-0.074)<br>*P* = 0.02 | 0.342<br>(-2.354-4.320)<br>*P* = 0.31 |
| Females | T2D<sub>PRS</sub> | PCOS | BMI<sub>residual</sub> | 0.268<br>(0.018-0.519)<br>*P* = 0.04 | 0.139<br>(0.046-0.242)<br>*P* = 0.002 | 0.338<br>(0.122-0.863)<br>*P* = 0.004 |

Lastly, the effects of BMI on PCOS comorbidities T2D, hypertension, and hypertensive heart disease were explored (**Table 5**). We found that BMI_residual_ was not a significant mediator for any of the tested models, even though clinical BMI was a significant mediator for T2D in females and a significant mediator for hypertensive heart disease in males. This again implicated *genetically regulated* BMI as the main source of the mediating effect for males since median BMI significantly mediated 14.8% of the variance in hypertensive heart disease (p = <2e-16) and mediated 23.7% of the variance in hypertension (p = 0.002) due to PCOS_PRS_. The lack of an association with BMI_resiudal_ in females also suggests *genetically regulated* BMI as the primary mediator for T2D, where it mediated 31.5% of the variance explained by the PCOS_PRS_.

**Table 5.** Genetically Regulated BMI is a Stronger Mediator in PCOS_PRS_ Shared Metabolic Pathways.

| Sex | Mediator | Outcome | ADE<br>$\beta$ (95% CI)<br>*P* | ACME<br>$\beta$ (95% CI)<br>*P* | Proportion<br>Mediated<br>(95% CI)<br>*P* |
| --- | --- | --- | --- | --- | --- |
| Sex-Combined | BMI | T2D | 0.009<br>(0.004-0.014)<br>*P* = <2e-16 | 0.005<br>(0.004-0.007)<br>*P* = <2e-16 | 0.367<br>(0.250 - 0.585)<br>*P* = <2e-16 |
|  | BMI<br>Residuals |  | 0.0134<br>(0.008- 0.019)<br>*P* = <2e-16 | 0.001<br>(-1.76E-04- 0.002)<br>*P* = 0.09 | 0.072<br>(-0.011- 0.155)<br>*P* = 0.09 |
|  | BMI | Hypertension | 0.007<br>(-2.13e-04-0.013)<br>*P* = 0.062 | 0.005<br>(0.004-0.006)<br>*P* = <2e-16 | 0.435<br>(0.269-1.043)<br>*P* = <2e-16 |
|  | BMI<br>Residuals |  | 0.012<br>(0.005-0.0178)<br>*P* = 0.002 | 8.89e-04<br>(-3.13E-04-0.002)<br>*P* = 0.13 | 0.072<br>(-0.036-0.207)<br>*P* = 0.13 |
|  | BMI | Hypertensive<br>Heart Disease | 0.008<br>(0.003-0.013)<br>*P* = 0.006 | 0.002<br>(0.001-0.003)<br>*P* = <2e-16 | 0.237<br>(0.144-0.457)<br>*P* = 0.002 |
|  | BMI<br>Residuals |  | 0.009<br>(0.004-0.014)<br>*P* = 0.002 | 0.0006<br>(-0.0001-0.001)<br>*P* = 0.09 | 0.062<br>(-0.011-0.152)<br>*P* = 0.09 |
| Females | BMI | T2D | 0.012<br>(0.005-0.019)<br>*P* = <2e-16 | 0.005<br>(0.004-0.007)<br>*P* = <2e-16 | 0.315<br>(0.208-0.535)<br>*P* = <2e-16 |
|  | BMI<br>Residuals |  | 0.016<br>(0.009-0.023)<br>*P* = <2e-16 | 9.64e-04<br>(-6.00e-04-0.003)<br>*P* = 0.22 | 0.058<br>(-0.037-0.154)<br>*P* = 0.22 |
|  | BMI | Hypertension | 0.002<br>(-0.007-0.010)<br>*P* = 0.73 | 0.006<br>(0.004-0.007)<br>*P* = <2e-16 | 0.742<br>(-2.165-5.363)<br>*P* = 0.08 |
|  | BMI Residuals |  | 0.007<br>(-0.002-0.015)<br>*P* = 0.11 | 9.01e-04<br>(-7.86E-04-0.002)<br>*P* = 0.26 | 0.112<br>(-0.426-0.806)<br>*P* = 0.30 |
|  | BMI | Hypertensive Heart Disease | 0.003<br>(-0.002- 0.009)<br>*P* = 0.31 | 0.002<br>(0.001- 0.003)<br>*P* = <2e-16 | 0.378<br>(-1.602-3.278)<br>*P* = 0.082 |
|  | BMI Residuals |  | 0.005<br>(-9.79e-04-0.011)<br>*P* = 0.11 | 5.50e-04<br>(-0.0002-0.001)<br>*P* = 0.14 | 0.097<br>(-0.367-0.781)<br>*P* = 0.20 |
| Males | BMI | T2D | 0.007<br>(-0.002-0.016)<br>*P* = 0.13 | 0.005<br>(0.003-0.007)<br>*P* = <2e-16 | 0.430<br>(0.186-1.377)<br>*P* = 0.02 |
|  | BMI Residuals |  | 0.011<br>(0.001-0.020)<br>*P* = 0.03 | 0.001<br>(-8.26e-04-0.003)<br>*P* = 0.27 | 0.086<br>(-0.115-0.421)<br>*P* = 0.28 |
|  | BMI | Hypertension | 0.014<br>(0.004-0.024)<br>*P* = 0.006 | 0.004<br>(0.003-0.006)<br>*P* = <2e-16 | 0.237<br>(0.127-0.530)<br>*P* = 0.002 |
|  | BMI Residuals |  | 0.018<br>(0.009-0.027)<br>*P* = <2e-16 | 8.65e-04<br>(-8.84e-04-0.003)<br>*P* = 0.30 | 0.046<br>(-0.049 -0.156)<br>*P* = 0.30 |
|  | BMI | Hypertensive Heart Disease | 0.015<br>(0.006- 0.025)<br>*P* = <2e-16 | 0.003<br>(0.001- 0.004)<br>*P* = <2e-16 | 0.148<br>(0.059-0.349)<br>*P* = <2e-16 |
|  | BMI Residuals |  | 0.017<br>(0.008- 0.027)<br>*P* = <2e-16 | 0.0006<br>(-8.82e-04- 0.002)<br>*P* = 0.43 | 0.034<br>(-0.063-0.139)<br>*P* = 0.43 |

PCOS<sub>PRS</sub> was analyzed as the exposure variable. *P* < 0.05 was considered statistically significant. ADE = average direct effect,
ACME = average causal mediation effect, CI = confidence interval

## Discussion

Through a comprehensive analysis of PCOS genetic risk across multiple phenotypes, we identified sex differences in the cardiometabolic traits associated with PCOS genetic risk. Among these, the most notable difference was that males with high PCOS_PRS_ were at greater risk of cardiovascular conditions than females at the same level of genetic risk. Genetically predicted BMI was revealed as a primary mediator of this risk in males, whereas both environmental and genetic BMI variance components mediated risk of T2D conferred by PCOS_PRS_ in females. Furthermore, only T2D_PRS_ and BMI_PRS_ were associated with PCOS diagnosis, indicating that many of the associations observed in the PCOS_PRS_ PheWAS were primarily driven by PCOS genetic risk and not the genetic effects of the identified comorbidities.

There is growing interest in studying PCOS related effects in males whether through their relationship to first-degree family members with PCOS or by an equivalent phenotype [34–36]. Generally, males with mothers or sisters with PCOS tend to exhibit a poorer cardiometabolic profile which can be observed as early as infancy [35]. A previous study suggests that males who exhibit a high genetic risk for PCOS are more likely to present with morbid obesity, T2D, and diabetes mellitus, two findings which were confirmed in this study [12]. Although we also found associations with CVD phenotypes in males, these were largely influenced by genetically predicted BMI which can significantly increase the lifetime risk for CVD and mortality rates of high-risk individuals [37]. Genetic susceptibility for PCOS could be an additional catalyst for these events in males, making individuals already predisposed to adverse metabolic outcomes more vulnerable.

In an effort to determine whether genetic risk for phenotypes comorbid with PCOS could also increase risk for PCOS, we conducted separate multivariable logistic regressions with PRS for BMI, diastolic blood pressure, systolic blood pressure, pulse pressure, T2D, heart failure, and CAD on the PCOS diagnosis outcome and found no significant associations outside of T2D_PRS_ and BMI_PRS_. This was unsurprising as the relationship between PCOS and CVD are still poorly elucidated and often debated [38]. It is possible that CVD is more prevalent in genetically high-risk males than females. The sex-difference in CVD prevalence may also be reflected in the PRS which may fail to fully capture CVD genetic risk for PCOS when stratified by sex due to fewer females in the discovery sample.

This study provides novel insight into PCOS genetic etiology and continues to underscore the importance of BMI in PCOS risk. As with T2D, we found that genetically predicted BMI is primarily responsible for the phenotypic association between T2D_PRS_ and PCOS. However, PCOS_PRS_ genetic risk led to more comorbid traits in the PheWAS in which both environmental and genetically predicted BMI influenced the metabolic associations. This may mean that PCOS patients with a family history of T2D can have an increased risk for morbid outcomes, as females with PCOS are already more likely to develop T2D at an earlier age [39]. This genetic susceptibility could also explain the high prevalence of insulin resistance in PCOS patients, which can be as high as 70% across all BMI strata [40]. These effects should be investigated further, as the genetic and biological pathways could differ in lean PCOS patients who also experience a high rate of insulin resistance [41].

This study offers many strengths. Firstly, we showed that cardiometabolic associations vary with sex and that the metabolic outcomes related to PCOS genetic architecture can be further understood by studying both males and females. Secondly, we decomposed BMI into genetically predicted (i.e., BMI_PRS_) and environmentally enriched (i.e., BMI_residual_) and evaluated their respective roles in mediating the cardiometabolic profiles associated with PCOS_PRS_. However, limitations include low power to detect any significant associations in our African descent sample. To date, there is no PCOS GWAS of African descent individuals, limiting all current similar studies to building PRS using European-based genetic variants, which do not perform as well in non-European populations [11,12]. Second, we only examined one environmental risk factor. Although many effects such as lifestyle and diet can be captured through BMI, it is not an exhaustive measurement nor does it accurately account for the full wellness of an individual [42,43]. Although other anthropometric features like hip-to-waist ratio (WHR) may be better indicators of health for some phenotypes [44], this information is not routinely collected in clinical settings or reported in EHRs. Furthermore, evidence does suggest that clinically ascertained BMI may be more informative for PCOS than WHR [45]. Finally, despite using the largest PCOS GWAS to date for this analysis, our PCOS_PRS_ still only explains a small portion of PCOS genetic variance. As these analyses expand, so too will our ability to detect the full genetic spectrum of PCOS and its subphenotypes.

PCOS is a multifaceted disorder with genetic architecture that is reflective of its heterogeneous outcomes. This polygenic structure captures a spectrum of metabolic comorbidities that is even more apparent when compared between sexes. Our findings show that males with high PCOS liability are indeed a high-risk group and those with a family history of PCOS should be closely monitored for hypertension and CVD. This is also true for females with PCOS and a family history of T2D, whose genetic risk could induce more severe comorbid outcomes. As such, management and screening strategies should be updated to reflect advances in PCOS etiology. This call to action is paramount and requires both widespread dissemination of risk factor information to relevant stakeholders and increases in PCOS research priorities and funding. This becomes even more crucial as PCOS comorbidities are often under-recognized in clinical settings and metabolic conditions are underutilized in PCOS screening methods [46–49].

## Data Availability

All GWAS summary statistics used in this study are publicly available and can be obtained from the following consortium groups and websites: International PCOS Consortium (https://doi.org/10.17863/CAM.27720), UK Biobank (http://www.nealelab.is/uk-biobank), HERMES (http://www.broadcvdi.org/), and CARDIoGRAMplusC4D (http://www.cardiogramplusc4d.org/). The summary statistics used from the Million Veteran Project can be accessed through dbGaP under accession number phs001672.v6.p1. The electronic health record data that support the findings of this study are available from Vanderbilt University Medical Center, but restrictions apply to the availability of these data, which were used under license for the current study, and so are not publicly available. Data are, however, available from the authors upon reasonable request and with permission of Vanderbilt University Medical Center.

## References

1. Rotterdam ESHRE/ASRM-Sponsored PCOS consensus workshop group. Revised 2003 consensus on diagnostic criteria and long-term health risks related to polycystic ovary syndrome (PCOS). Hum Reprod. 2004;19: 41–47. doi:10.1093/humrep/deh098

2. Teede HJ, Misso ML, Costello MF, Dokras A, Laven J, Moran L, et al. Recommendations from the international evidence-based guideline for the assessment and management of polycystic ovary syndrome. Hum Reprod. 2018;33: 1602–1618. doi:10.1093/humrep/dey256

3. Shorakae S, Boyle J, Teede H. Polycystic ovary syndrome: a common hormonal condition with major metabolic sequelae that physicians should know about. Intern Med J. 2014;44: 720–726. doi:10.1111/imj.12495

4. Gibson-Helm M, Teede H, Dunaif A, Dokras A. Delayed Diagnosis and a Lack of Information Associated With Dissatisfaction in Women With Polycystic Ovary Syndrome. J Clin Endocrinol Metab. 2017;102: 604–612. doi:10.1210/jc.2016-2963

5. March WA, Moore VM, Willson KJ, Phillips DIW, Norman RJ, Davies MJ. The prevalence of polycystic ovary syndrome in a community sample assessed under contrasting diagnostic criteria. Human Reproduction. 2010;25: 544–551. doi:10.1093/humrep/dep399

6. Cussons AJ, Stuckey BGA, Walsh JP, Burke V, Norman RJ. Polycystic ovarian syndrome: marked differences between endocrinologists and gynaecologists in diagnosis and management. Clin Endocrinol. 2005;62: 289–295. doi:10.1111/j.1365-2265.2004.02208.x

7. Azziz R, Carmina E, Dewailly D, Diamanti-Kandarakis E, Escobar-Morreale HF, Futterweit W, et al. Criteria for Defining Polycystic Ovary Syndrome as a Predominantly Hyperandrogenic Syndrome: An Androgen Excess Society Guideline. The Journal of Clinical Endocrinology & Metabolism. 2006;91: 4237–4245. doi:10.1210/jc.2006-0178

8. Sanchez-Garrido MA, Tena-Sempere M. Metabolic dysfunction in polycystic ovary syndrome: Pathogenic role of androgen excess and potential therapeutic strategies. Mol Metab. 2020;35: 100937. doi:10.1016/j.molmet.2020.01.001

9. Dapas M, Lin FTJ, Nadkarni GN, Sisk R, Legro RS, Urbanek M, et al. Distinct subtypes of polycystic ovary syndrome with novel genetic associations: An unsupervised, phenotypic clustering analysis. PLoS Med. 2020;17: e1003132. doi:10.1371/journal.pmed.1003132

10. Zhang Y, Movva VC, Williams MS, Lee MTM. Polycystic Ovary Syndrome Susceptibility Loci Inform Disease Etiological Heterogeneity. JCM. 2021;10: 2688. doi:10.3390/jcm10122688

11. Actkins KV, Singh K, Hucks D, Velez Edwards DR, Aldrich M, Cha J, et al. Characterizing the clinical and genetic spectrum of polycystic ovary syndrome in electronic health records. J Clin Endocrinol Metab. 2020. doi:10.1210/clinem/dgaa675

12. Joo YY, Actkins K, Pacheco JA, Basile AO, Carroll R, Crosslin DR, et al. A Polygenic and Phenotypic Risk Prediction for Polycystic Ovary Syndrome Evaluated by Phenome-Wide Association Studies. J Clin Endocrinol Metab. 2020;105: 1918–1936. doi:10.1210/clinem/dgz326

13. Lee H, Oh J-Y, Sung Y-A, Chung HW. A genetic risk score is associated with polycystic ovary syndrome-related traits. Hum Reprod. 2016;31: 209–215. doi:10.1093/humrep/dev282

14. Vink JM, Sadrzadeh S, Lambalk CB, Boomsma DI. Heritability of polycystic ovary syndrome in a Dutch twin-family study. J Clin Endocrinol Metab. 2006;91: 2100–2104. doi:10.1210/jc.2005-1494

15. Day FR, Hinds DA, Tung JY, Stolk L, Styrkarsdottir U, Saxena R, et al. Causal mechanisms and balancing selection inferred from genetic associations with polycystic ovary syndrome. Nat Commun. 2015;6: 8464. doi:10.1038/ncomms9464

16. Day F, Karaderi T, Jones MR, Meun C, He C, Drong A, et al. Large-scale genome-wide meta-analysis of polycystic ovary syndrome suggests shared genetic architecture for different diagnosis criteria. PLoS Genet. 2018;14: e1007813. doi:10.1371/journal.pgen.1007813

17. Louwers YV, Lao O, Fauser BCJM, Kayser M, Laven JSE. The impact of self-reported ethnicity versus genetic ancestry on phenotypic characteristics of polycystic ovary syndrome (PCOS). J Clin Endocrinol Metab. 2014;99: E2107–16. doi:10.1210/jc.2014-1084

18. Bjonnes AC, Saxena R, Welt CK. Relationship between polycystic ovary syndrome and ancestry in European Americans. Fertil Steril. 2016;106: 1772–1777. doi:10.1016/j.fertnstert.2016.08.033

19. Dapas M, Sisk R, Legro RS, Urbanek M, Dunaif A, Hayes MG. Family-based quantitative trait meta-analysis implicates rare noncoding variants in DENND1A in polycystic ovary syndrome. J Clin Endocrinol Metab. 2019. doi:10.1210/jc.2018-02496

20. Robinson JR, Wei W-Q, Roden DM, Denny JC. Defining Phenotypes from Clinical Data to Drive Genomic Research. Annu Rev Biomed Data Sci. 2018;1: 69–92. doi:10.1146/annurev-biodatasci-080917-013335

21. Bien SA, Wojcik GL, Zubair N, Gignoux CR, Martin AR, Kocarnik JM, et al. Strategies for Enriching Variant Coverage in Candidate Disease Loci on a Multiethnic Genotyping Array. PLoS One. 2016;11: e0167758. doi:10.1371/journal.pone.0167758

22. Abraham G, Qiu Y, Inouye M. FlashPCA2: principal component analysis of Biobank-scale genotype datasets. Bioinformatics. 2017;33: 2776–2778. doi:10.1093/bioinformatics/btx299

23. Dennis JK, Sealock JM, Straub P, Lee YH, Hucks D, Actkins K, et al. Clinical laboratory test-wide association scan of polygenic scores identifies biomarkers of complex disease. Genome Med. 2021;13: 6. doi:10.1186/s13073-020-00820-8

24. Locke AE, Kahali B, Berndt SI, Justice AE, Pers TH, Day FR, et al. Genetic studies of body mass index yield new insights for obesity biology. Nature. 2015;518: 197–206. doi:10.1038/nature14177

25. Gaziano JM, Concato J, Brophy M, Fiore L, Pyarajan S, Breeling J, et al. Million Veteran Program: A mega-biobank to study genetic influences on health and disease. J Clin Epidemiol. 2016;70: 214–223. doi:10.1016/j.jclinepi.2015.09.016

26. Giri A, Hellwege JN, Keaton JM, Park J, Qiu C, Warren HR, et al. Trans-ethnic association study of blood pressure determinants in over 750,000 individuals. Nat Genet. 2019;51: 51–62. doi:10.1038/s41588-018-0303-9

27. Vujkovic M, Keaton JM, Lynch JA, Miller DR, Zhou J, Tcheandjieu C, et al. Discovery of 318 new risk loci for type 2 diabetes and related vascular outcomes among 1.4 million participants in a multi-ancestry meta-analysis. Nat Genet. 2020;52: 680–691. doi:10.1038/s41588-020-0637-y

28. Shah S, Henry A, Roselli C, Lin H, Sveinbjörnsson G, Fatemifar G, et al. Genome-wide association and Mendelian randomisation analysis provide insights into the pathogenesis of heart failure. Nat Commun. 2020;11: 163. doi:10.1038/s41467-019-13690-5

29. Munz M, Richter GM, Loos BG, Jepsen S, Divaris K, Offenbacher S, et al. Genome-wide association meta-analysis of coronary artery disease and periodontitis reveals a novel shared risk locus. Sci Rep. 2018;8: 13678. doi:10.1038/s41598-018-31980-8

30. Ge T, Chen C-Y, Ni Y, Feng Y-CA, Smoller JW. Polygenic prediction via Bayesian regression and continuous shrinkage priors. Nat Commun. 2019;10: 1776. doi:10.1038/s41467-019-09718-5

31. Barber TM, Hanson P, Weickert MO, Franks S. Obesity and Polycystic Ovary Syndrome: Implications for Pathogenesis and Novel Management Strategies. Clin Med Insights Reprod Health. 2019;13: 1179558119874042. doi:10.1177/1179558119874042

32. Bulik-Sullivan B, Finucane HK, Anttila V, Gusev A, Day FR, Loh P-R, et al. An atlas of genetic correlations across human diseases and traits. Nat Genet. 2015;47: 1236–1241. doi:10.1038/ng.3406

33. Tingley D, Yamamoto T, Hirose K, Keele L, Imai K. Mediation: R package for causal mediation analysis. J Stat Softw. 2014. Available: https://oar.princeton.edu/jspui/handle/88435/pr1gj2f

34. Di Guardo F, Ciotta L, Monteleone M, Palumbo M. Male Equivalent Polycystic Ovarian Syndrome: Hormonal, Metabolic, and Clinical Aspects. Int J Fertil Steril. 2020;14: 79–83. doi:10.22074/ijfs.2020.6092

35. Recabarren SE, Smith R, Rios R, Maliqueo M, Echiburú B, Codner E, et al. Metabolic profile in sons of women with polycystic ovary syndrome. J Clin Endocrinol Metab. 2008;93: 1820–1826. doi:10.1210/jc.2007-2256

36. Subramaniam K, Tripathi A, Dabadghao P. Familial clustering of metabolic phenotype in brothers of women with polycystic ovary syndrome. Gynecol Endocrinol. 2019;35: 601–603. doi:10.1080/09513590.2019.1566451

37. Khan SS, Ning H, Wilkins JT, Allen N, Carnethon M, Berry JD, et al. Association of Body Mass Index With Lifetime Risk of Cardiovascular Disease and Compression of Morbidity. JAMA Cardiol. 2018;3: 280–287. doi:10.1001/jamacardio.2018.0022

38. Papadakis G, Kandaraki E, Papalou O, Vryonidou A, Diamanti-Kandarakis E. Is cardiovascular risk in women with PCOS a real risk? Current insights. Minerva Endocrinol. 2017;42: 340–355. doi:10.23736/S0391-1977.17.02609-8

39. Rubin KH, Glintborg D, Nybo M, Abrahamsen B, Andersen M. Development and Risk Factors of Type 2 Diabetes in a Nationwide Population of Women With Polycystic Ovary Syndrome. J Clin Endocrinol Metab. 2017;102: 3848–3857. doi:10.1210/jc.2017-01354

40. Diamanti-Kandarakis E, Dunaif A. Insulin resistance and the polycystic ovary syndrome revisited: an update on mechanisms and implications. Endocr Rev. 2012;33: 981–1030. doi:10.1210/er.2011-1034

41. Escobar-Morreale HF. Polycystic ovary syndrome: definition, aetiology, diagnosis and treatment. Nat Rev Endocrinol. 2018;14: 270–284. doi:10.1038/nrendo.2018.24

42. Rothman KJ. BMI-related errors in the measurement of obesity. Int J Obes (Lond). 2008;32 Suppl 3: S56–59. doi:10.1038/ijo.2008.87

43. Chrysant SG, Chrysant GS. The single use of body mass index for the obesity paradox is misleading and should be used in conjunction with other obesity indices. Postgrad Med. 2019;131: 96–102. doi:10.1080/00325481.2019.1568019

44. Li C, Engström G, Hedblad B, Calling S, Berglund G, Janzon L. Sex differences in the relationships between BMI, WHR and incidence of cardiovascular disease: a population-based cohort study. Int J Obes (Lond). 2006;30: 1775–1781. doi:10.1038/sj.ijo.0803339

45. Venkatesh SS, Ferreira T, Benonisdottir S, Rahmioglu N, Becker CM, Granne I, et al. The role of obesity in female reproductive conditions: A Mendelian Randomisation study. Genetic and Genomic Medicine; 2021 Jun. doi:10.1101/2021.06.01.21257781

46. Rodgers RJ, Avery JC, Moore VM, Davies MJ, Azziz R, Stener-Victorin E, et al. Complex diseases and co-morbidities: polycystic ovary syndrome and type 2 diabetes mellitus. Endocr Connect. 2019;8: R71–R75. doi:10.1530/EC-18-0502

47. Piltonen TT, Ruokojärvi M, Karro H, Kujanpää L, Morin-Papunen L, Tapanainen JS, et al. Awareness of polycystic ovary syndrome among obstetrician-gynecologists and endocrinologists in Northern Europe. PLOS ONE. 2019;14: e0226074. doi:10.1371/journal.pone.0226074

48. Dokras A, Saini S, Gibson-Helm M, Schulkin J, Cooney L, Teede H. Gaps in knowledge among physicians regarding diagnostic criteria and management of polycystic ovary syndrome. Fertil Steril. 2017;107: 1380-1386.e1. doi:10.1016/j.fertnstert.2017.04.011

49. Mott MM, Kitos NR, Coviello AD. Practice Patterns in Screening for Metabolic Disease in Women with PCOS of Diverse Race-Ethnic Backgrounds. Endocr Pract. 2014;20: 855–863. doi:10.4158/EP13414.OR

