## Supplementary Figures for "Sex Modifies the Effect of Genetic Risk Scores for Polycystic Ovary Syndrome on Metabolic Phenotypes"

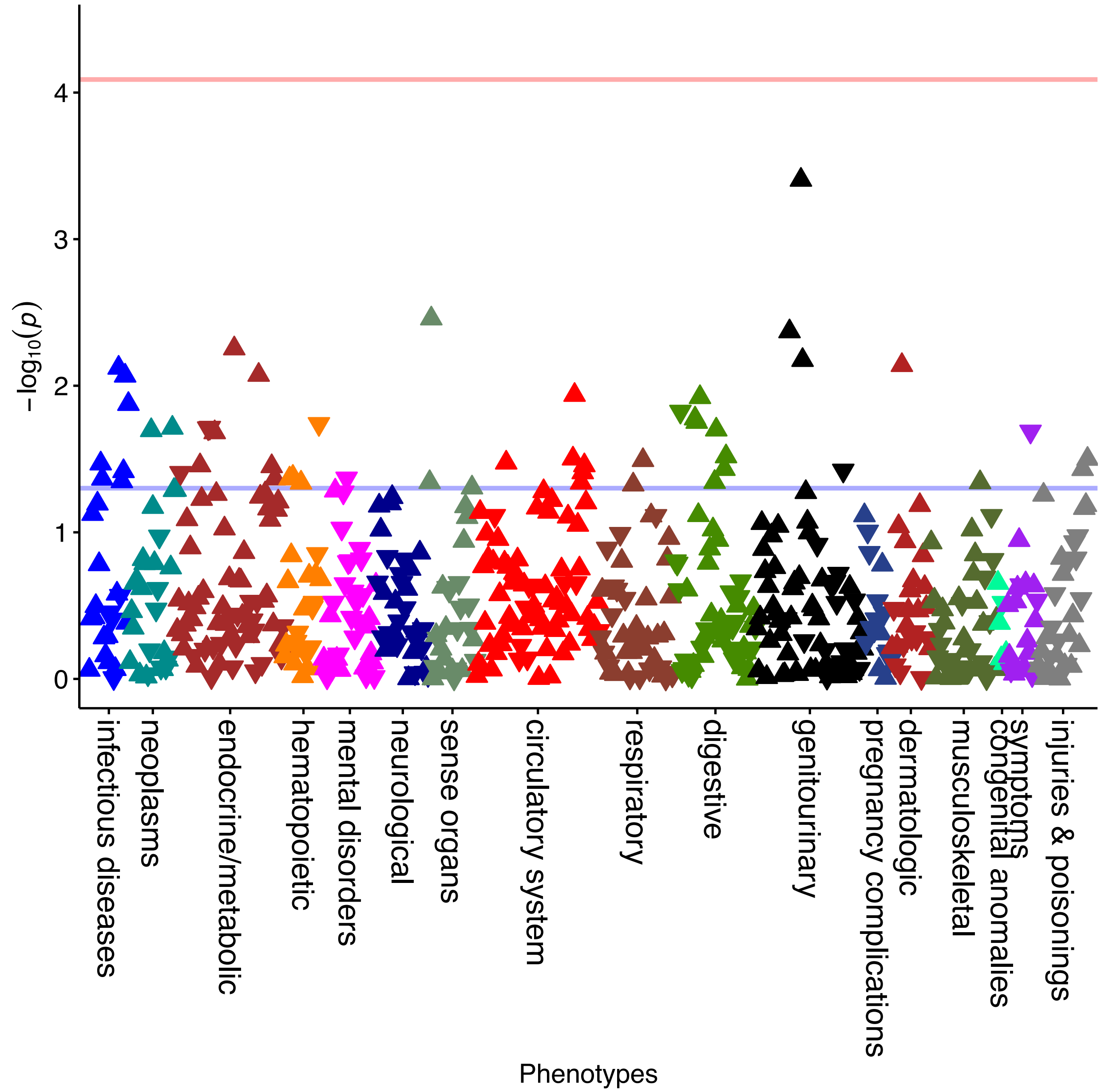


**Supplementary Figure 1. The African Descent $PCOS_{PRS}$ PheWAS is Underpowered.** No associations passed Bonferroni correction (P= 8.14e-05) for $PCOS_{PRS}$ calculated in African ancestry individuals. The red line represents the Bonferroni correction and the blue represents the false discovery rate of $P < 0.05$.

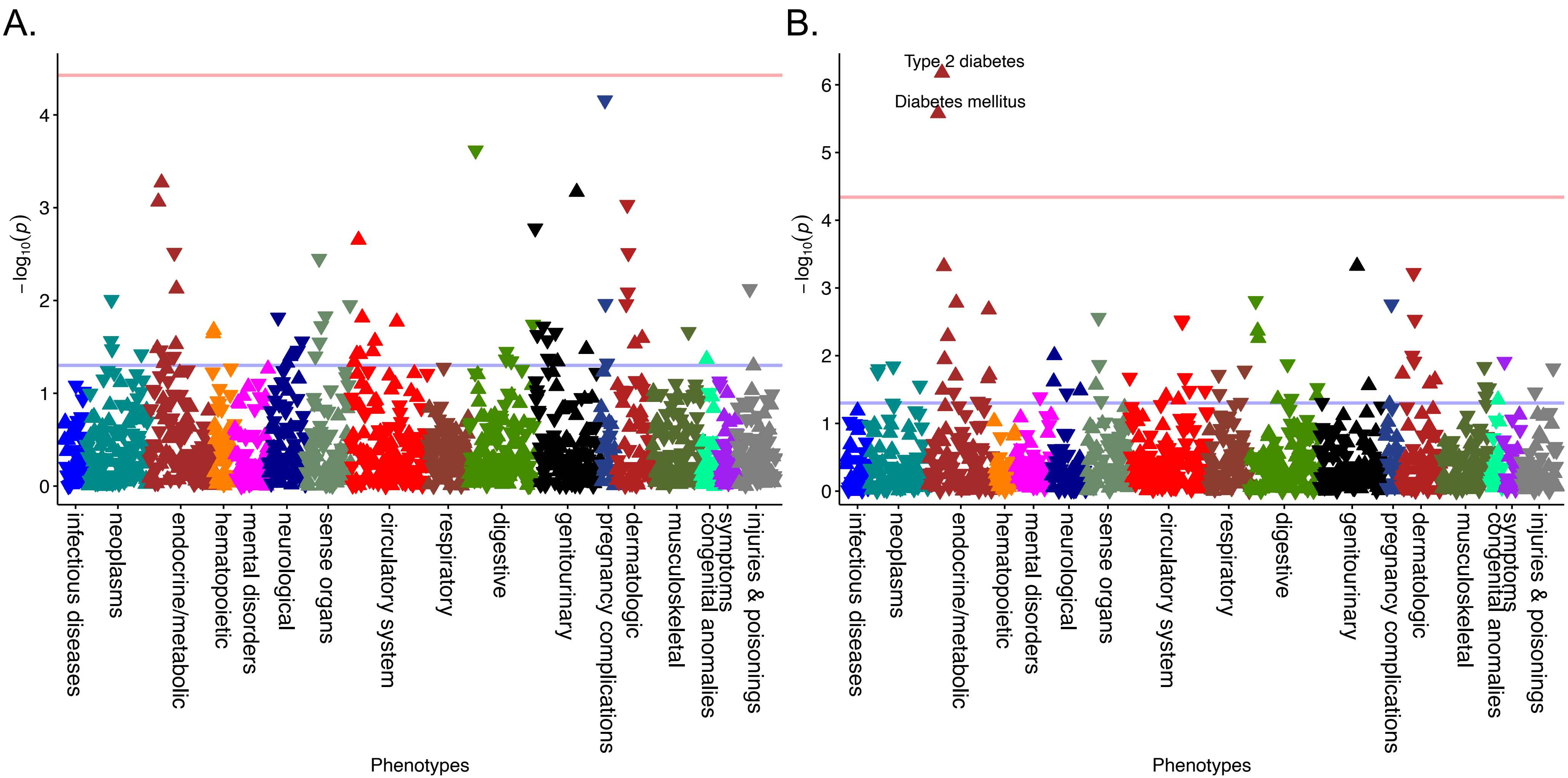


**Supplementary Figure 2. Pleiotropic Associations are Influenced by Phenotypic Features of PCOS.** To determine the robustness of the observed significant associations with $PCOS_{PRS}$, sensitivity analyses were employed. First, (A) $PCOS_{PRS}$ was adjusted for BMI in everyone. No associations passed Bonferroni correction, represented by the red line (P=3.74e-05). The blue line represents a false discovery rate of 0.05. To identify what phenotypes were not the result of a diagnosis, (B) females were stratified and further adjusted for PCOS case status (P=4.57e-05e).

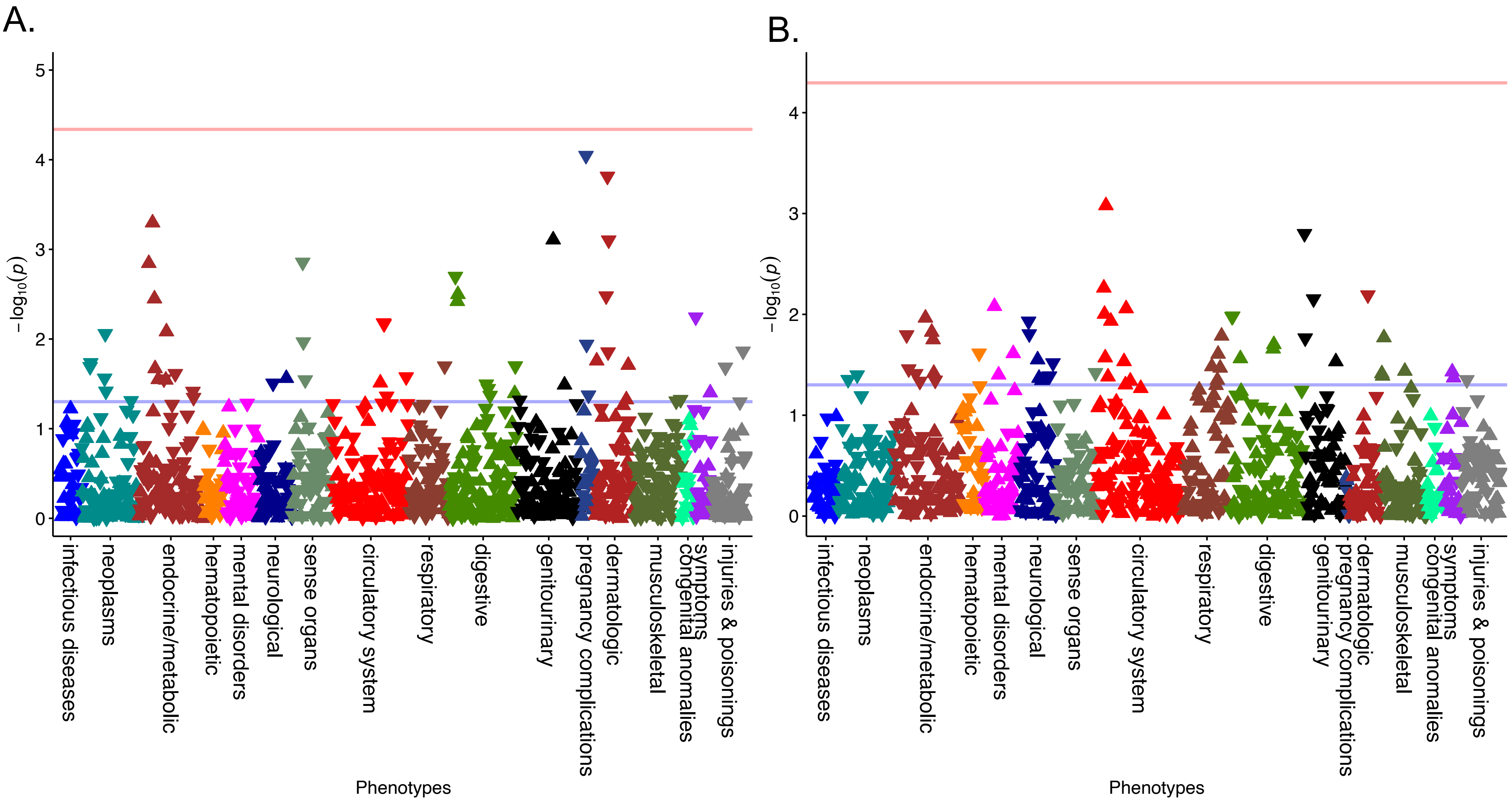


**Supplementary Figure 3. Associations Driven by Sex Differentiated PCOS$_{PRS}$ Can Be Explained by BMI.** Sex stratified analyses were adjusted for body mass index (BMI) in a sensitivity analysis. Results are shown for (A) females (P=5.07e-05) and (B) males (P=4.90e-05). The red line represents the Bonferroni correction and the blue represents the false discovery rate of P < 0.05.

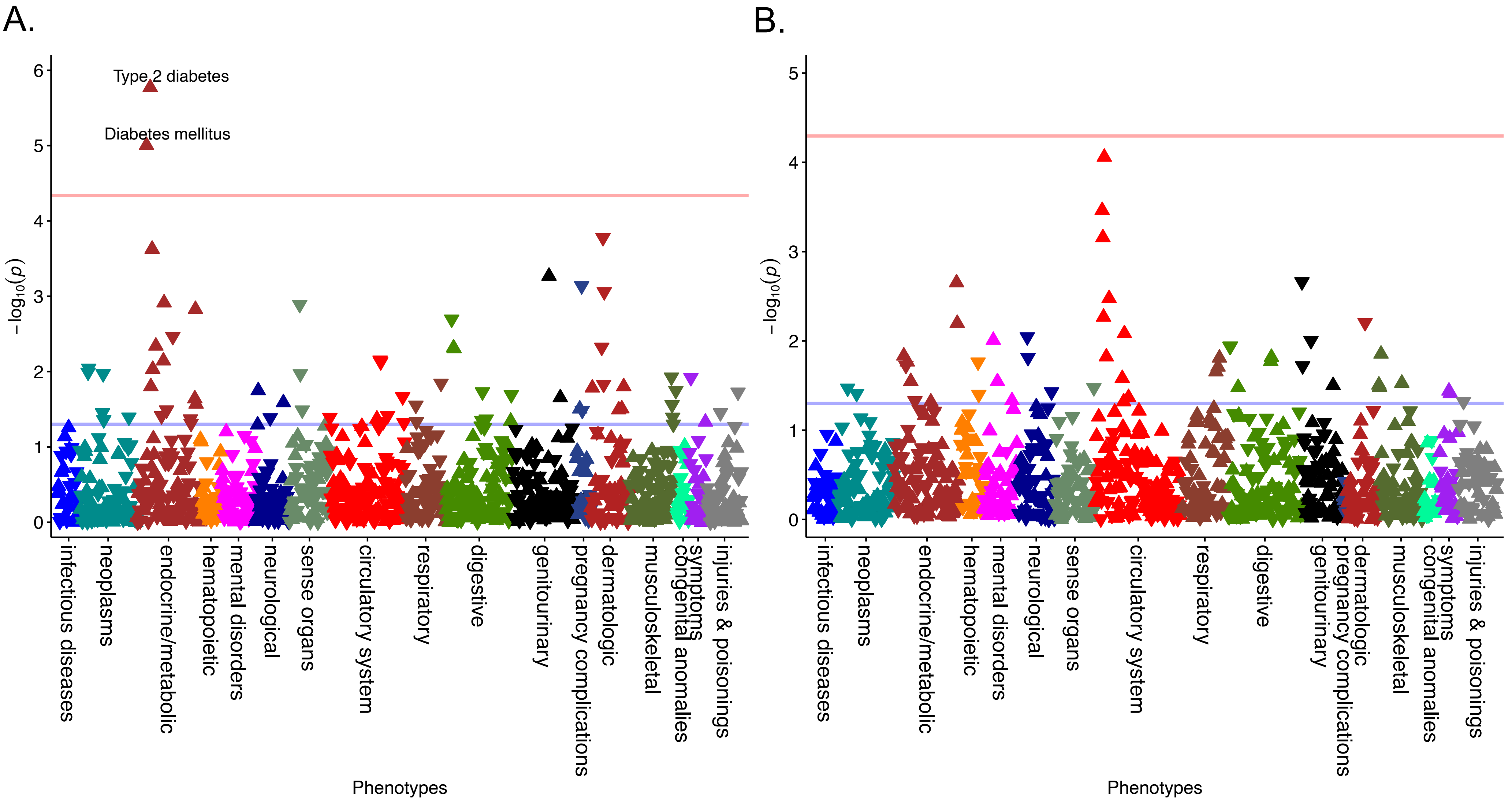


**Supplementary Figure 4. Accounting for BMI$_{residual}$ in the PCOS$_{PRS}$ PheWAS Analysis Improves Associations.** PCOS$_{PRS}$ were stratified by sex and adjusted for BMI$_{residual}$ in addition to age and the top ten principal components for (A) females and (B) males. The Bonferroni correction for females was P=4.59e-05 and P=5.07e-05 for males. The red line represents the Bonferroni correction and the blue represents the false discovery rate of $P < 0.05$.

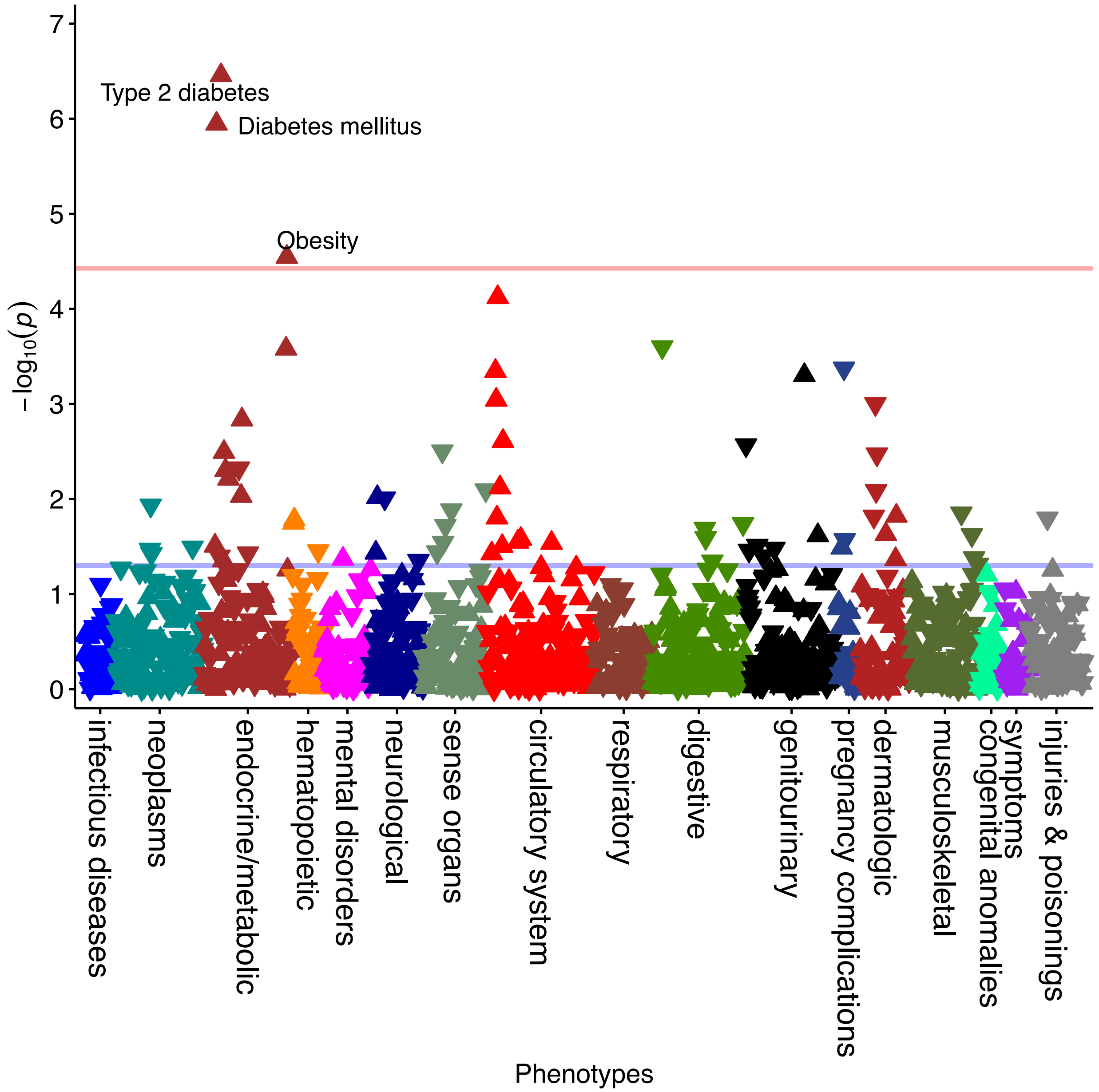


**Supplementary Figure 5. Accounting for $BMI_{residual}$ Improves Detection of Obesity in the Sex-Combined Dataset.** The model was adjusted for adjusted for $BMI_{residual}$, age, and the top ten principal components. The red line represents the Bonferroni correction of P = 3.74e-05 and the blue represents the false discovery rate of P < 0.05.
